# Cost-effectiveness of High-Dose Influenza Vaccination in the Netherlands: Updated Analysis Incorporating New Evidence

**DOI:** 10.64898/2026.02.17.26346451

**Authors:** Simon van der Pol, Sajad Emamipour, Anita van Oudheusden, Bas Slierendregt, Gerald Moncayo, Cornelis Boersma

**Affiliations:** Health-Ecore, Zeist, the Netherlands; University of Groningen, University Medical Center Groningen, Department of Health Sciences, Groningen, the Netherlands; Sanofi Netherlands, Amsterdam, The Netherlands; Health Economics and Value Assessment, Sanofi, Lyon, France; Open University, Department of Management Sciences, Heerlen, the Netherlands

**Keywords:** High-dose inactivated influenza vaccination, Standard-dose inactivated influenza vaccination, Cardiorespiratory hospitalizations Cost-effectiveness analysis

## Abstract

**Background:** High-dose inactivated influenza vaccination (HD-IIV) demonstrates superior effectiveness versus standard-dose vaccination (SD-IIV) in adults aged ≥60 years. A recent meta-analysis integrated complementary evidence sources of representing over 85 million individuals across 14 influenza seasons.

**Methods:** A previously developed model was updated using life-time horizon and societal perspective. Updated parameters included demographics, costs, hospitalization rates, and relative vaccine effectiveness (rVE): RCT evidence (24% for ILI, 7% for cardiorespiratory hospitalizations) and RCT + real-world evidence (RWE) (15% for ILI, 8% for cardiorespiratory hospitalizations).

**Results:** HD-IIV resulted in incremental cost-effectiveness ratios of €7,300/QALY (RCT evidence) and €5,800/QALY (RCT+RWE evidence). Implementation would prevent 7,200 general practitioner visits, 6,300 cardiorespiratory hospitalizations, and 269 deaths, by using RCT evidence. Probabilistic sensitivity analysis demonstrated >99% probability of cost-effectiveness at €20,000/QALY threshold for both RCT and RCT+RWE evidence.

**Conclusions:** HD-IIV remains highly cost-effective for Dutch adults aged ≥60 years under updated evidence scenarios, supporting implementation in the national immunization programme.

**Highlights:**

- The economic analysis of high-dose inactivated influenza vaccine was updated.
- Relative vaccine effectiveness of HD-IIV incorporating recent evidence was used.
- HD-IIV remains cost-effective in Dutch adults aged ≥60.

## Introduction

Each year, influenza forms a major burden on healthcare systems worldwide. Vaccination programmes aim to reduce the burden on both the patient and the system levels. Recent innovations in influenza vaccines aim to enhance the protection against influenza [1]. These vaccines mainly focus on older adults who are at increased risk due to age-related immunosenescence and the increased prevalence of comorbidities [1]. High-dose inactivated influenza vaccination (HD-IIV) is registered for adults aged 60 and over in the Netherlands. This vaccine has shown superior effectiveness against influenza cases and hospitalizations against standard dose, both in randomized clinical trials (RCTs) and real-world evidence (RWE) [2].

Currently, enhanced influenza vaccines have not been implemented in the Netherlands. An advice by the Health Council dated December 2024, concluded that potentially two vaccines can be implemented in the Netherlands for adults aged 60 and over, in the existing immunization programme, if proven to be cost-effective [1]. In 2024, Van der Pol et al. showed that HD-IIV can be considered to be cost-effective compared to SD-IIV, with a base-case incremental cost-effectiveness ratio (ICER) of 5,400 per quality-adjusted life year (QALY) gained considering RCT evidence and €300 per QALY considering RCT+RWE [3].

A recent pooled analysis of two methodologically harmonized, individually randomized trials (FLUNITY-HD) involving 466,320 individuals in Denmark and Spain demonstrated that HD-IIV provides statistically significant effectiveness against influenza-related hospitalizations and extends to cardiorespiratory and all-cause hospitalizations in adults aged 65+ years across the 2022-2025 seasons compared to SD-IIV. The authors report an adjusted relative vaccine effectiveness (rVE) of 6.3% against cardiorespiratory hospitalization and 31.9% against laboratory-confirmed influenza hospitalization [4].

To provide a comprehensive assessment of HD-IIV effectiveness, an updated meta-analysis was conducted including both RCT and RWE from >85 million individuals aged ≥65 years across 15 influenza seasons up to the 2024/25 season [2]. The updated meta-analysis demonstrated that HD-IIV provided statistically significant greater protection than SD-IIV against a broad range of outcomes, including probable or laboratory-confirmed influenza-like illness (ILI), influenza-related hospitalizations, respiratory, cardiovascular, cardiorespiratory, and all-cause hospitalizations, as well as influenza and pneumonia-related emergency room visits or hospitalizations [2].

Considering the effectiveness against cardio-respiratory hospitalizations was the most influential parameter on the cost-effectiveness of HD-IIV in the Netherlands [3], we believe it is important to update the previous analysis using these recent meta-analysis findings showing rVE of 15% for probable or laboratory-confirmed ILI and 8% for cardiorespiratory hospitalizations [2]. In this paper, we present an updated analysis of switching from SD-IIV to HD-IIV in the Netherlands, using rVE measures from both RCT and RCT+RWE. Where relevant, other inputs are also updated to reflect recent data, such as costs and demographics.

## Material and methods

### Model description

The model was unchanged compared to the previous published economic analysis [3], simulating two strategies for the Netherlands: SD-IIV, which is the current standard of care, and the introduction of HD-IIV. Figure 1 shows a graphical representation of the decision-tree model which can be used to model the occurrence of influenza, including potential GP consultations, and hospitalizations, potentially followed by in-hospital mortality. One influenza season was considered in the model, mortality and productivity losses were incorporated from a lifetime horizon and productivity losses were included adhering to Dutch guidelines for economic evaluations. The main outcome considered was the ICER expressed in euros per QALY. In addition to QALYs, life years (LYs) were also reported. LYs and QALYs were discounted with 1.5% annually.

**Figure 1.**
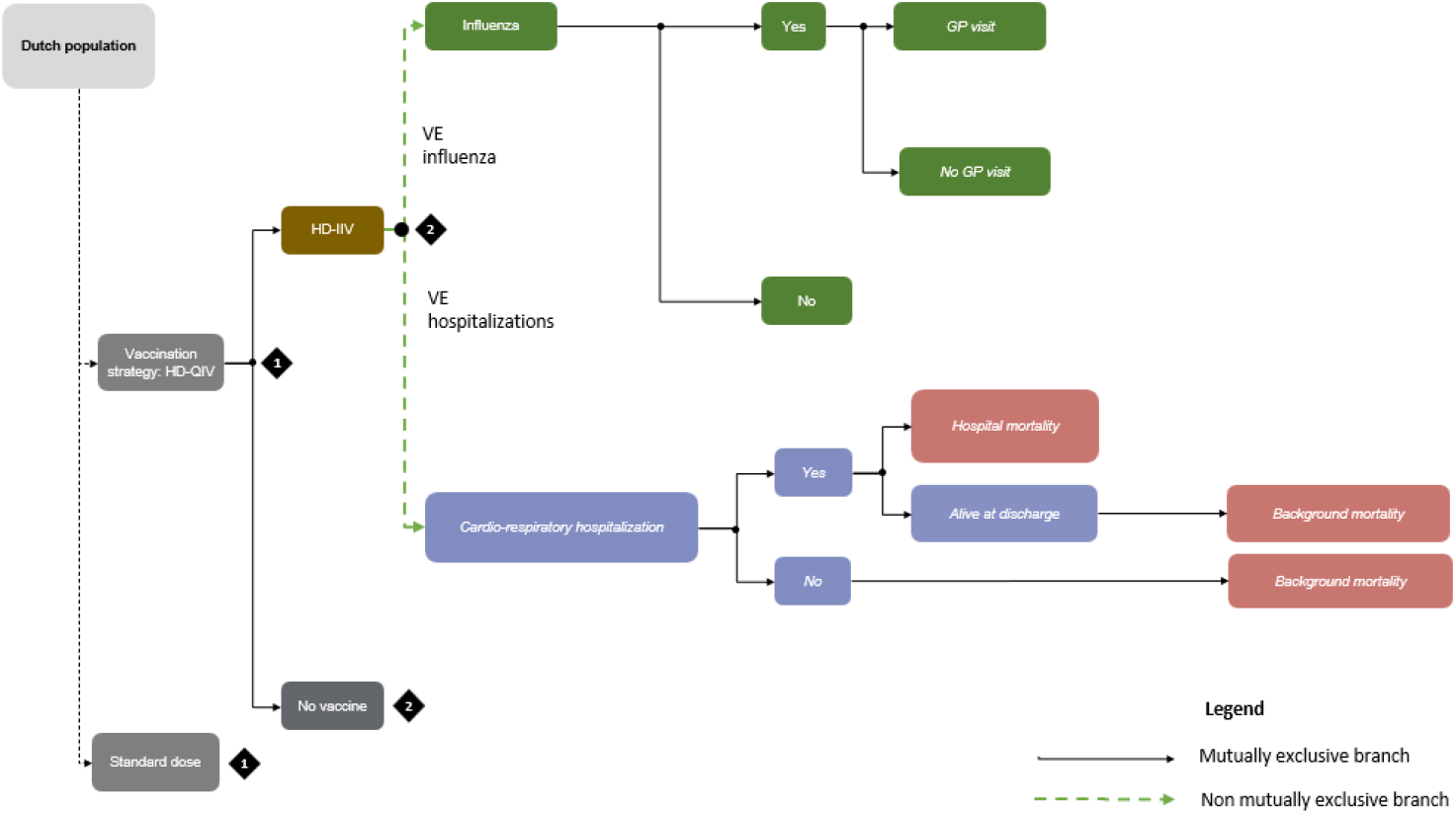
graphical representation the of influenza health-economic model. HD-IIV: high-dose inactivated influenza vaccination, GP: general practitioner, VE: vaccine efficacy

### Updated input parameters

Table 1 provides a summary of the updated clinical inputs for the model, including the distributions and ranges used for the probabilistic sensitivity analysis (PSA). All other inputs are included in the supplementary materials.

**Table 1.** updated parameters.

| Parameter | Value (range) | Distribution | Reference |
| --- | --- | --- | --- |
| **Epidemiological and demographic input** |  |  |  |
| Vaccination coverage | Age 60-64, low-risk: 34.2%<br>Age 60-64, high-risk: 51.5%<br>Age 65-74, low-risk: 57.5%<br>Age 65-74, high-risk: 67.8%<br>Age 75+, low-risk: 69.9%<br>Age 75+, high-risk: 76.6% | None | [6] |
| Eligible population | 4,879,198 |  | [5] |
| **Healthcare use** |  |  |  |
| Hospitalization rate<br>cardiorespiratory causes<br>per 100,000 population | Age 60-64: 1,546 (1,160-1,933)<br>Age 65-74: 4,666 (3,500 – 5,833)<br>Age 75+: 6,695 (5,021 – 8,368) | Beta | [7] |
| **Vaccine efficacy** |  |  |  |
| Relative vaccine<br>efficacy HD-IIV versus<br>SD-IIV, influenza-like<br>illness | RCT+RWE estimate: 15% (5% - 24%)<br>RCT estimate: 24% (10% - 36%) | Lognormal | [2] |
| Relative vaccine efficacy HD-IIV versus SD-IIV, cardiorespiratory hospitalizations | RCT+RWE estimate: 8% (5% - 11%)<br>RCT estimate: 7% (4% - 10%) | Lognormal | [2] |
| **Outcomes** |  |  |  |
| Mortality rate following cardiorespiratory hospitalization | Age 60-64: 1.09% (0.82-1.37)<br>Age 65-74: 2.34% (1.76-2.93)<br>Age 75+: 5.82% (4.37-7.28) | Beta | [8] |
| **Costs** |  |  |  |
| Vaccine administration costs | €14.13 |  | [11] |
| Influenza-related visit general practitioner | €34.21 |  | [12] |
| Hospitalization costs, per cardiorespiratory episode | €6,697 |  | [13,14] |
| Average daily wages | €354 |  | [15] |
| Workforce participation rate | Age 60-64: 68.7%<br>Age 65 – 74: 19.1%<br>Age 75+: 0% |  | [16]<br>[16]<br>Assumption |
| Friction period | 125 days |  | [10] |

*HD-IIV: high-dose inactivated influenza vaccination; QALY: Quality-adjusted life year; RCT: Randomized clinical trial; RWE: Real-world evidence, based on retrospective cohort studies and test-negative control studies; SD-IIV: standard-dose inactivated influenza vaccination*

In the update, demographic data and vaccine coverage were updated [5,6]. Hospitalization rates and subsequent mortality from cardiorespiratory hospitalizations were derived from the Statistics Netherlands for the year 2023[7,8].

#### Comparative vaccine effectiveness

We updated the analyses using RCT and RCT+RWE evidence from a recent meta-analysis [2]. Using RCT+RWE, the rVE changed to 15% against ILI and 8% against cardiorespiratory hospitalizations [2]. We also incorporated a scenario using RCT evidence only, with a rVE of 24% against ILI and 7% against cardiorespiratory hospitalizations [2].

#### Costs

The vaccine unit costs were not changed compared to the previous analysis, €8 for SD-IIV and €32 for HD-IIV. All other costs were inflated to the 2025 price levels using the consumer price index of the Statistics Netherlands [9]. The friction period, relevant for productivity losses, was updated to a period of 125 days [10]. Since no costs beyond the scope of one influenza season were included in the analysis, no discounting was applied to costs.

### Scenario and sensitivity analyses

Deterministic (DSA) and probabilistic sensitivity analyses (PSA) were performed to assess the uncertainty of parameters in the model. The DSA was reported using a tornado diagram showing the 10 most influential parameters and the PSA consisting of 1,000 simulations was visualized using a cost-effectiveness plane and a cost-effectiveness acceptability curve (CEAC). In line with the previous analysis, willingness-to-pay thresholds of €20,000 and €50,000 per QALY were included. The scenario analyses remained the same and parameters were updated.

## Results

Implementing HD-IIV would result in preventing 7,200 GP visits and over 6,000 hospitalizations. HD-IIV led to an incremental 2,458 QALYs, 2,529 life years and 229,198 saved days absent from work. The cost-effectiveness results are displayed in table 2. Incorporating the updated RWE yields an ICER of €5,800 per QALY, up from €300 per QALY. For the updated scenario where only RCT evidence is considered, HD-IIV is cost-effective with an ICER of €7,300 per QALY, up from €5,400 per QALY. The disaggregated costs and clinical outcomes are displayed in the supplementary materials (tables 4 and 5). The scenario analyses are displayed in supplementary tables 6 – 10. The deterministic sensitivity analysis is displayed in supplementary figure 1.

**Table 2.**
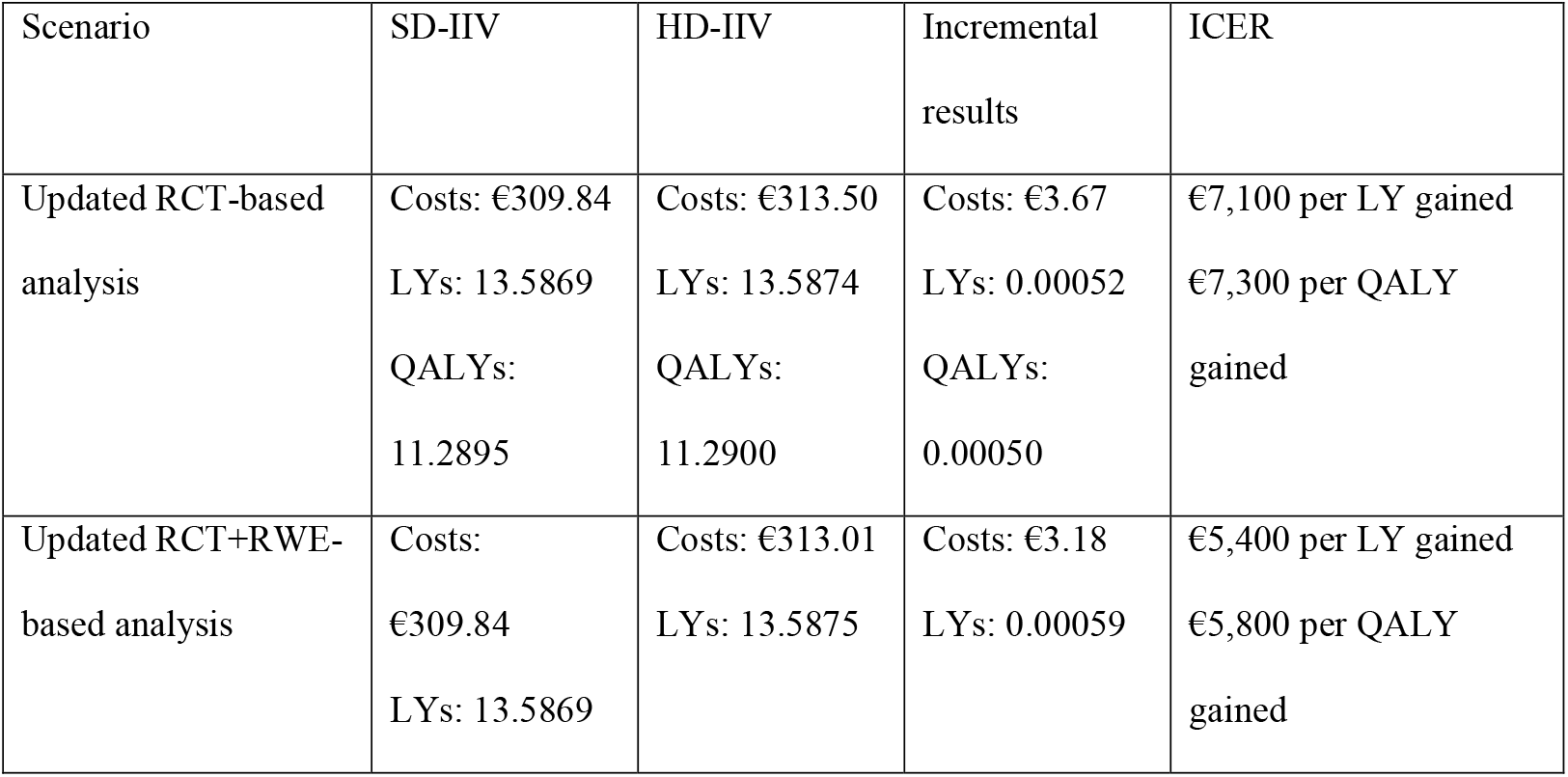
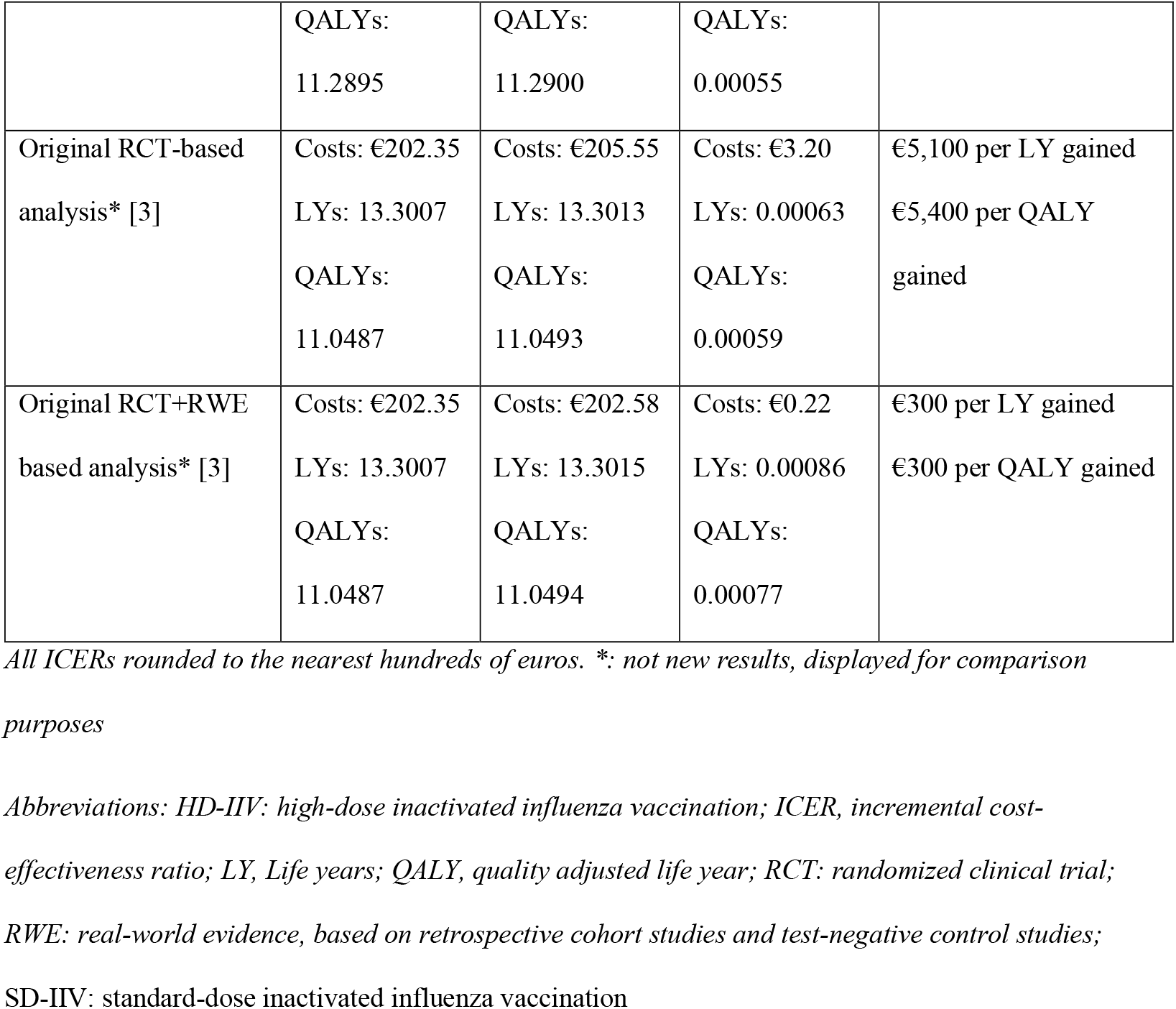
Base case results and scenario analyses, ICERs are rounded to the nearest hundreds of euros.

### Probabilistic sensitivity analysis

Figure 2 displays the CEAC for both the RCT and the RCT+RWE analysis. HD-IIV can be considered cost-effective in over 99% of the simulations at the WTP of €20,000 per QALY. The CE plane is included in supplementary figure 2.

**Figure 2.**
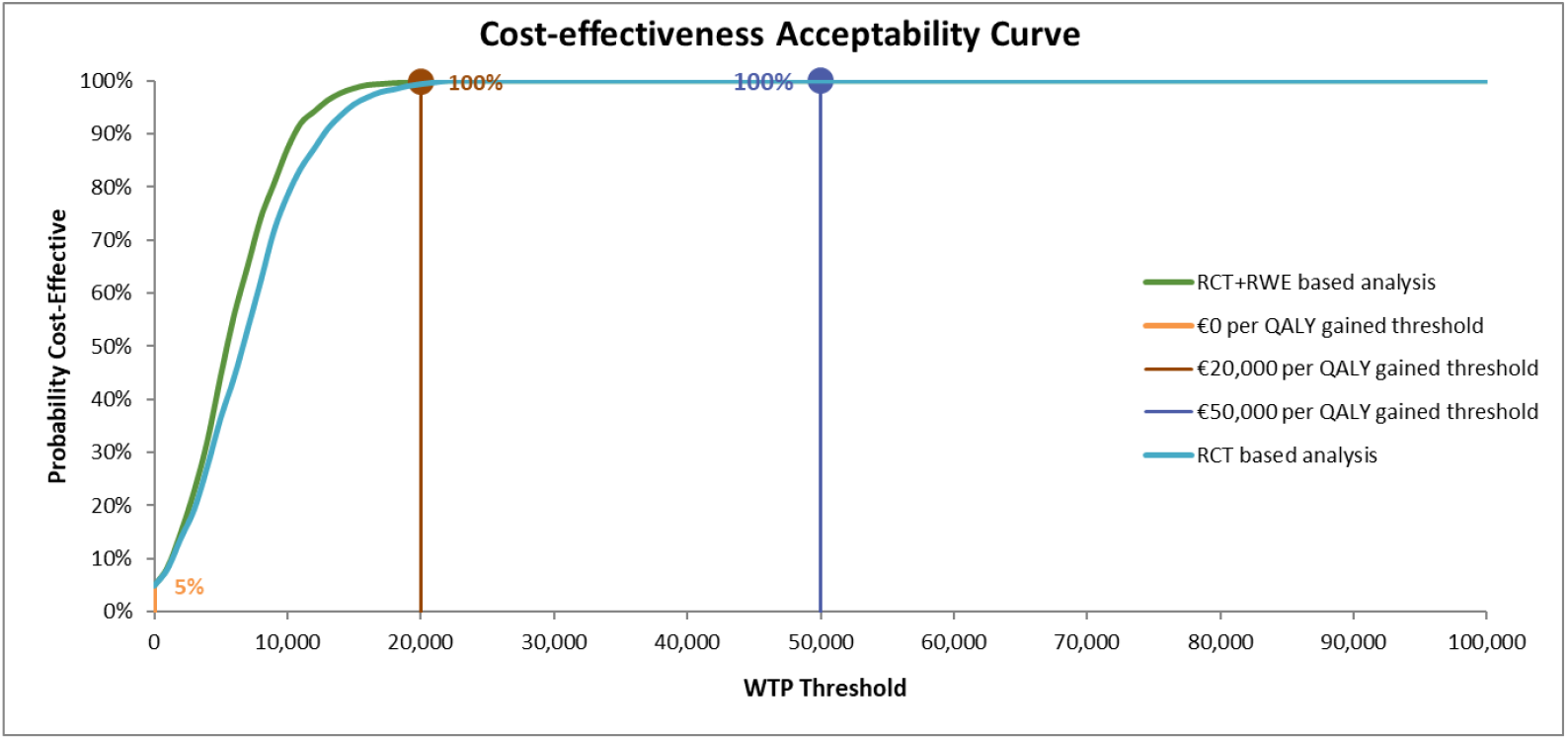
Cost-effectiveness acceptability curve for different WTP threshold and incorporating for the RCT and RCT+RWE analyses. RCT: Randomized Clinical Trial; RWE: Real-World Evidence; WTP: willingness to pay; QALY: quality-adjusted life year

## Discussion

In our updated analysis, we find that the major conclusions regarding the cost-effectiveness of HD-IIV in the Netherlands remain unchanged. Incorporating the recent evidence on HD-IIV as well as prior evidence [2,4], the ICER increased compared to our previous analyses since the effectiveness of HD-IIV compared to SD-IIV was reduced [3]. The rVE of HD-IIV compared with SD-IIV against cardiorespiratory hospitalizations decreased from 12.2% to 7% in the updated meta-analysis based on RCT evidence. The updated ICER is €7,300 per QALY for the RCT analysis and €5,800 per QALY for the RCT+RWE analysis. Nevertheless, HD-IIV is still a highly cost-effective option to improve the health of the ageing population in the Netherlands and to alleviate the burden on the healthcare system during Winter. The conclusion that HD-IIV is cost-effective is consistent across all included scenarios.

The reduction in effectiveness of HD-IIV against cardiorespiratory hospitalization may be explained by the increased uptake of COVID-19 and pneumococcal vaccines, which may also have an impact on this outcome. Coverage of COVID-19 and pneumococcal vaccines were high in FLUNITY-HD (71% and 96%, respectively) in both the HD-IIV and SD-IIV arms, which may contribute to the reduction of the overall risk of cardiorespiratory disease. Two main effects dampen the increasing trend on the ICER caused by the incorporation of the FLUNITY-HD study. First, compared to the 2024 analysis for the Netherlands, the total QALYs lost due to hospitalizations have increased due to a higher reported incidence of cardiorespiratory hospitalizations [3,7]. Second, costs were inflated to the levels of 2025 which increases the overall costs. The costs for SD-IIV and HD-IIV remained the same, favouring the implementation of HD-IIV.

In their advice of 2024, the Dutch Health Council lamented the absence of comparative evidence between HD-IIV and its closest competitor, the adjuvanted influenza vaccine (aIV) [1]. We previously concluded that the comparative evidence between the adjuvanted and high-dose vaccines is severely limited, as no head-to-head randomized trials have been performed and that high-quality comparative clinical evidence was required to compare HD-IIV to aIV [3,17]. Unfortunately, this evidence still is not available, and we were not able to include a meaningful comparison between the two competing vaccines. Currently, a RCT of SD-IIV compared to aIV is in progress, which may yield the evidence required to indirectly compare HD-IIV and aIV [18].

Few limitations remain from our previous analysis. In this analysis, nursing home mortality was not considered. Additionally, we did not incorporate the long-term care required after discharge from influenza-related hospitalizations. A recent US study reported that 15.8% of patients hospitalized for influenza required subsequent long-term care [19]. Although comparable data for the Netherlands are currently unavailable, incorporating those parameters would likely further improve the cost-effectiveness of HD-IIV. Also, many studies included adults aged 65 and over [2], while a minimum age of 60 is more relevant for the Dutch context.

The cost-effectiveness of vaccines will always be closely linked to the transmission of infectious diseases through the populations, which can be difficult to predict. In some seasons, the influenza vaccine will be more effective, and cost-effective, than in other seasons. Therefore, it is important to incorporate the most comprehensive data from as many seasons as possible, including both the robustness of RCTs and the broader generalizability of RWE, to ensure cost-effectiveness estimates reflect the variability inherent across influenza seasons. Frequent monitoring of the incidence and effectiveness is important to keep decision-making up-to-date and optimize the protection of vulnerable populations from communicable diseases.

## Conclusions

This analysis incorporated updated estimates for the effectiveness of HD-IIV, as well as more recent incidence of cardio-respiratory hospitalizations. Based on various parameter assumptions, HD-IIV remains cost-effective considering the most recent evidence. Considering the appraisal of the Dutch Health Council [1], HD-IIV yields a valid option to be included in the Dutch immunization programme for older adults.

## Author contributions

All authors attest they meet the ICMJE criteria for authorship.

Conceptualization: SvdP, AvO, BS, CB; Data curation: SvdP, SE; Formal analysis: SE; Funding acquisition: SvdP, CB; Investigation: SvdP, SE, GM; Methodology: SvdP, SE, AvO, BS, GM, CB; Project administration: SvdP; Resources: SvdP, CB; Supervision: CB; Validation: SvdP, AvO, BS, GM; Visualization: SE; Writing – original draft: SvdP, SE; Writing – review & editing: AvO, BS, GM, CB

## Acknowledgements

We would like to thank Chris de Jong from Sanofi for his input and support during the writing of the paper. We thank Sanofi for supplying the model which was adapted to run the analyses detailed in this paper.

## Data statement

Data and materials are available upon reasonable request.

