## Supplementary materials for "Cost-effectiveness of High-Dose Influenza Vaccination in the Netherlands: Updated Analysis Incorporating New Evidence"

### Supplementary table 1: overview of model parameters

| Parameter | Value (range) | Distribution | Reference |
|---|---|---|---|
| **Epidemiological and demographic input** | | | |
| Influenza attack rate unvaccinated cohort | 7.2% (4.3-12) | Uniform | (1) |
| Vaccination coverage | Age 60-64, low-risk: 34.2%<br>Age 60-64, high-risk: 51.5%<br>Age 65-74, low-risk: 57.5%<br>Age 65-74, high-risk: 67.8%<br>Age 75+, low-risk: 69.9%<br>Age 75+, high-risk: 76.6% | None | (2) |
| Proportion of influenza A and B | A: 68% (31-99)<br>B: 32% (1-69) | Beta | (3) |
| Eligible population | 4,879,198 | | (4) |
| **Healthcare use** | | | |
| Probability of general practitioner visit | Age 60-64: 20.1% (15.1-25.1)<br>Age 65+: 29.6% (22.2-37.0) | Gamma | (5,6) |
| Hospitalization rate cardiorespiratory causes per 100,000 population | Age 60-64: 1,546 (1,160-1,933)<br>Age 65-74: 4,666 (3,500 – 5,833)<br>Age 75+: 6,695 (5,021 – 8,368) | Beta | (7) |
| Hospital length of stay | 5 days (3.8-6.3) | Gamma | (8) |
| Proportion of cardiorespiratory hospitalizations during the influenza season (October – April) | 68% (50-82) | Uniform | (9) |
| **Vaccine efficacy** | | | |
| Relative vaccine efficacy HD-IIV versus SD-IIV, influenza-like illness | RCT+RWE estimate: 15% (5% - 24%)<br>RCT estimate: 24% (10% - 36%) | Lognormal | [2] |
| Relative vaccine efficacy HD-IIV versus SD-IIV, cardiorespiratory hospitalizations | RCT+RWE estimate: 15% (5% - 24%)<br>RCT estimate: 7% (4% - 10%) | Lognormal | [2] |
| Relative vaccine efficacy standard-dose vaccine versus no vaccination, influenza-like illness | 50% (39%-65%) | Lognormal | (12,13) |

| | | | |
|---|---|---|---|
| Relative vaccine efficacy standard-dose vaccine versus no vaccination, cardiorespiratory hospitalizations | 28.5% (10.0%-42.3%) | Lognormal | (14,15) |
| **Outcomes** | | | |
| Mortality rate following cardiorespiratory hospitalization | Age 60-64: 1.09% (0.82-1.37)<br>Age 65-74: 2.34% (1.76-2.93)<br>Age 75+: 5.82% (4.37-7.28) | Beta | (16) |
| Duration of influenza episode | 7.73 days | None | (17) |
| Quality adjusted life days lost due to influenza-like illness (per day) | 0.32 | Gamma | (18,19) |
| Total QALYs lost due to cardiorespiratory hospitalization | 0.018 | Normal | (20) |

### Supplementary table 2: overview of costs

| Cost item | Value | Reference |
|---|---|---|
| Standard-dose vaccine | €8 | Estimation based on cost-effectiveness SD-QIV (5) |
| High-dose vaccine | €32 | List price |
| Vaccine administration costs | €14.13 | (21) |
| Influenza-related visit general practitioner | €34.21 | (24) |
| Hospitalization costs, per cardiorespiratory episode | €6,697 | (5,23) |
| Productivity losses | | |
| Average daily wages | €354 | (24) |
| Workforce participation rate | Age 60-64: 68.7%<br>Age 65 – 74: 19.1%<br>Age 75+: 0% | (25)<br>(25)<br>Assumption |
| Friction period | 125 days | (26) |

### Supplementary table 3: updated parameters for scenario with only respiratory complications

| Parameter | Value | Reference |
|---|---|---|
| **Hospitalization rate respiratory causes per 100,000 population** | Age 60-64: 379<br>Age 65-74: 1,345<br>Age 75+: 2,100 | (27) |
| **Hospital length of stay** | 5 days | (8) |
| **Proportion of respiratory hospitalizations during the influenza season** | 68% | (9) |
| **Mortality probability following respiratory hospitalization** | Age 60-64: 2.34%<br>Age 65-74: 4.59%<br>Age 75+: 8.93% | (16) |
| **Total QALYs lost due to respiratory hospitalization** | 0.018 | (20) |

### Supplementary table 4: disaggregated costs for the base case

| Costs | SD-IIV | | HD-IIV | | Difference | |
|---|---|---|---|---|---|---|
| | Original model (28) | Updated model | Original model (28) | Updated model | Original model (28) | Updated model |
| Vaccine costs | 20,308,123 € | 23,690,290 € | 81,232,494 € | 94,761,159 € | 60,924,370 € | 71,070,869 € |
| Vaccine Administration | 32,366,072 € | 41,838,928 € | 32,366,072 € | 41,838,928 € | 0 € | 0 € |
| General practitioner visit costs | 2,217,258 € | 2,180,896 € | 2,005,096 € | 2,006,298 € | -212,162 € | -245,727 € |
| Hospitalization costs | 652,232,161 € | 1,023,776,527 € | 611,077,393 € | 981,690,838 € | -41,154,769 € | -42,085,688 € |
| Productivity losses (hospitalizations) | 70,164,229 € | 156,022,885 € | 65,542,100 € | 145,503,812 € | -4,622,130 € | -10,519,073 € |
| Productivity losses (mortality) | 152,560,281 € | 264,174,730 € | 152,321,983 € | 263,839,345 € | -238,298 € | -335,385 € |
| **Total costs** | **929,848,125 €** | **1,511,755,384 €** | **944,545,137 €** | **1,529,640,380 €** | **14,697,012 €** | **17,884,996 €** |

### Supplementary table 5: disaggregated clinical outcomes for the base case

| Clinical Outcomes | SD-IIV | | HD-IIV | | Difference | |
|---|---|---|---|---|---|---|
| | Original model (28) | Updated model | Original model (28) | Updated model | Original model (28) | Updated model |
| Influenza Cases | 239,468 | 244,696 | 217,443 | 219,110 | -22,024 | -25,586 |
| Influenza-related GP Visits | 64,530 | 65,833 | 58,356 | 58,650 | -6,175 | -7,183 |
| Cardio-respiratory disease Hospitalization | 115,855 | 152,876 | 108,545 | 146,591 | -7,310 | -6,284 |
| Bed Occupancy (days) | 579,276 | 764,380 | 542,724 | 732,957 | -36,551 | -31,422 |
| Lost Work Days due to Influenza | 2,430,360 | 2,655,879 | 2,223,562 | 2,426,681 | -206,798 | -229,198 |
| Influenza-related mortality | 4,982 | 6,132 | 4,660 | 5,863 | -322 | -269 |
| Background Mortality | 110,095 | 106,650 | 110,108 | 106,660 | 13 | 10 |
| Total QALYs | 50,770,772 | 55,083,694 | 50,773,497 | 55,086,152 | 2,725 | 2,458 |
| Total LYs | 61,119,224 | 66,293,267 | 61,122,108 | 66,295,796 | 2,885 | 2,529 |

### Supplementary table 6: results for scenario with healthcare perspective, considering RCT evidence

| Model | Vaccine | Total costs (€) | Total Lys | Total QALYs | Difference costs (€) | Difference LYs | Difference QALYs | ICER (€/LY) | ICER (€/QALY) |
|---|---|---|---|---|---|---|---|---|---|
| Updated model | SD-IIV | 223.72 | 13.5869 | 11.2895 | -- | - | - | - | - |
| | HD-IIV | 229.61 | 13.5874 | 11.2900 | 5.89 | 0.000518 | 0.000504 | 11,363 | 11,694 |
| Original model (28) | SD-IIV | 153.88 | 13.3007 | 11.0487 | -- | - | - | - | - |
| | HD-IIV | 158.14 | 13.3013 | 11.0493 | 4.26 | 0.000628 | 0.000593 | 6,779 | 7,176 |

### Supplementary table 7: results for scenario of SD-IIV priced at €4 per dose, considering RCT evidence

| Model | Vaccine | Total costs (€) | Total Lys | Total QALYs | Difference costs (€) | Difference LYs | Difference QALYs | ICER (€/LY) | ICER (€/QALY) |
|---|---|---|---|---|---|---|---|---|---|
| Updated model | SD-IIV | 307.41 | 13.5869 | 11.2895 | - | - | - | - | - |
| | HD-IIV | 313.50 | 13.5874 | 11.2900 | 6.09 | 0.000518 | 0.000504 | 11,755 | 12,097 |
| Original model (28) | SD-IIV | 200.14 | 13.3007 | 11.0487 | - | - | - | - | - |
| | HD-IIV | 205.55 | 13.3013 | 11.0493 | 5.41 | 0.000628 | 0.000593 | 8,614 | 9,118 |

### Supplementary table 8: results for scenario of SD-IIV priced at €16.51 per dose, considering RCT evidence

| Model | Vaccine | Total costs (€) | Total Lys | Total QALYs | Difference costs (€) | Difference LYs | Difference QALYs | ICER (€/LY) | ICER (€/QALY) |
|---|---|---|---|---|---|---|---|---|---|
| Updated model | SD-IIV | 315.00 | 13.5869 | 11.2895 | - | - | - | - | - |
| | HD-IIV | 313.50 | 13.5878 | 11.2903 | -1.5 | 0.000903 | 0.000835 | HD-IIV dominates SD-IIV | HD-IIV dominates SD-IIV |
| Original model (28) | SD-IIV | 207.05 | 13.3007 | 11.0487 | - | - | - | - | - |
| | HD-IIV | 205.55 | 13.3013 | 11.0493 | -1.5 | 0.000628 | 0.000593 | HD-IIV dominates SD-IIV | HD-IIV dominates SD-IIV |

### Supplementary table 9: results for scenario of considering respiratory hospitalizations, instead of cardiorespiratory hospitalizations, considering RCT evidence

| Model | Vaccine | Total costs (€) | Total Lys | Total QALYs | Difference costs (€) | Difference LYs | Difference QALYs | ICER (€/LY) | ICER (€/QALY) |
|---|---|---|---|---|---|---|---|---|---|
| Updated model | SD-IIV | 162.69 | 13.5925 | 11.2944 | - | - | - | - | - |
| | HD-IIV | 167.95 | 13.5932 | 11.2951 | 5.25 | 0.000712 | 0.000658 | 7,378 | 7,982 |
| Original model (28) | SD-IIV | 106.09 | 13.3051 | 11.0525 | - | - | - | - | - |
| | HD-IIV | 113.52 | 13.3057 | 11.0531 | 7.43 | 0.000548 | 0.000517 | 13,544 | 14,352 |

Supplementary table 10: results for scenario of considering respiratory hospitalizations, instead of cardiorespiratory hospitalizations, considering RCT+RWE evidence (Relative vaccine efficacy HD-IIV versus SD-IIV, influenza-like illness: 15%, relative vaccine efficacy HD-IIV versus SD-IIV, respiratory hospitalizations: 12%)

| Model | Vaccine | Total costs (€) | Total Lys | Total QALYs | Difference costs (€) | Difference LYs | Difference QALYs | ICER (€/LY) | ICER (€/QALY) |
|---|---|---|---|---|---|---|---|---|---|
| Updated model | SD-IIV | 162.69 | 13.5925 | 11.2944 | - | - | - | - | - |
| | HD-IIV | 171.14 | 13.5930 | 11.2949 | 8.44 | 0.0004748 | 0.0004365 | 17,785 | 19,348 |

### Supplementary figure 1: Tornado diagram

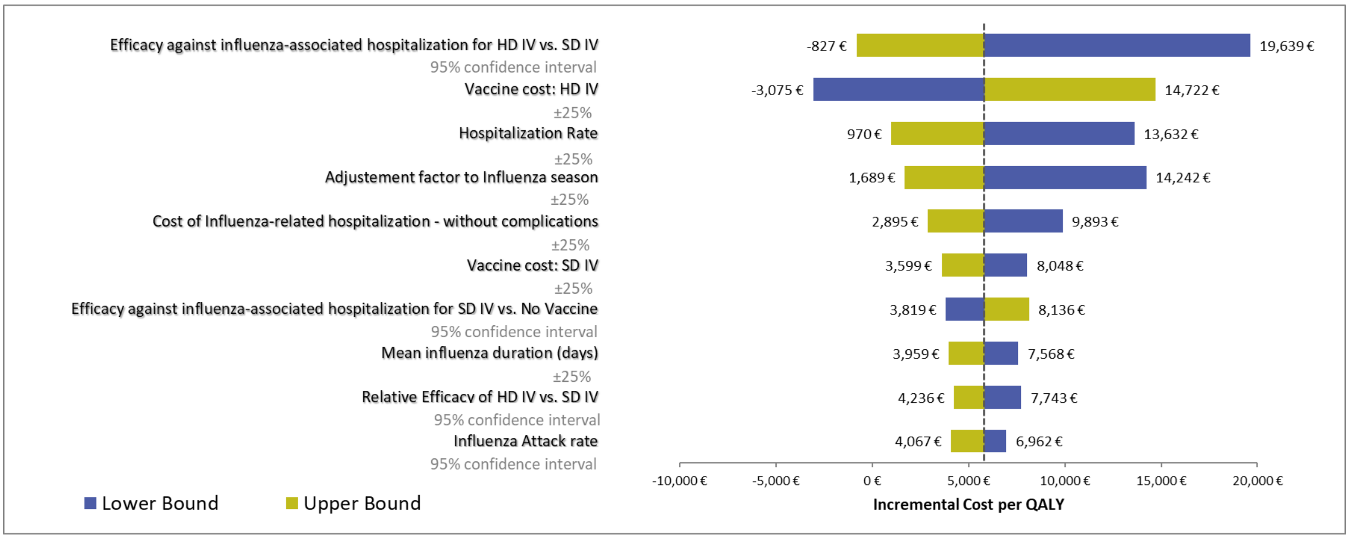


*Tornado diagram of deterministic sensitivity analysis based on RCT+RWE analysis, all negative ICERs are in the South-West Quadrant, i.e., HD-IV is cost-saving HD: High Dose, SD: Standard Dose; IV: Influenza Vaccine.*

### Supplementary figure 2: cost-effectiveness plane

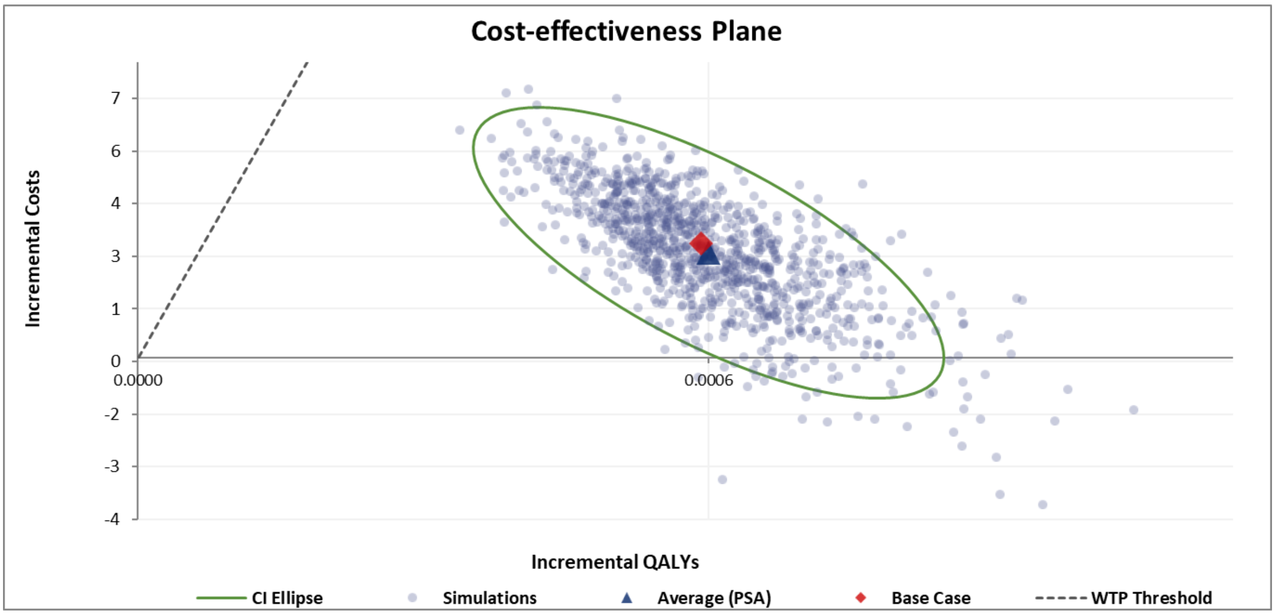


*Cost-effectiveness planes for the RCT+RWE analysis, the displayed WTP threshold is €20.000 per QALY, PSA: probabilistic sensitivity analysis, WTP: willingness to pay, QALY: quality-adjusted life year*

11. LEE UPDATE PLACEHOLDER.

22. Hakkaart - van Roijen L, Tan SS, Bouwmans CAM. Handleiding voor Kostenonderzoek. Diemen: College van Zorgverzekeringen; 2010.
